# Reliability and Validity of Cognitive Workload in Older Adults

**DOI:** 10.1101/2020.10.26.20219881

**Authors:** Hannes Devos, Kathleen Gustafson, Pedram Ahmadnezhad, Ke Liao, Jonathan D. Mahnken, William M. Brooks, Jeffrey M. Burns

## Abstract

Cognitive workload (mental effort) is a measure of attention allocation to a task, which can be administered through self-report or physiological measures. Cognitive workload is increasingly recognized as an important determinant of performance in cognitive tests and daily life activities. However, the reliability and validity of these measures have not been established in older adults with a wide range of cognitive ability. The aim of this study was to establish the test-retest reliability of the NASA-Task Load Index (NASA-TLX) and Index of Cognitive Activity (ICA), extracted from pupillary size. The convergent validity of these measures against event-related potentials (ERPs) was also investigated. A total of 38 individuals with scores on the Montreal Cognitive Assessment ranging between 17 and 30 completed a working memory test (n-back) with three levels of difficulty at baseline and two-week follow-up. Intraclass correlation coefficients (ICC) values of the NASA-TLX ranged between 0.71 and 0.81, demonstrating good to excellent reliability. Mean ICA scores showed fair to good reliability, with ICC’s ranging between 0.56 and 0.73. Mean ICA and NASA-TLX scores showed significant and moderate correlations (Pearson r range between 0.30 and 0.33) with P3 ERP at the midline channels. We conclude that ICC and NASA-TLX are reliable measures of cognitive workload in older adults. Further research is needed in dissecting the subjective and objective constructs of cognitive workload.

## 1 Introduction

Despite its incredible power and flexibility, there are limits to the brain’s capabilities. For example, working memory — the storage space that provides the foundation for higher-order cognitive functions— is on average only able to retain four items at any given time.[1] Performance on working memory tests is determined by the brain’s ability to allocate attention (mental effort) to the task and its available resources. Mental effort has been conceptualized as “cognitive workload” and if measured well may have properties that offer relevant information beyond that provided by standard performance measures such as accuracy or response times of a task. Cognitive workload has traditionally been characterized as a direct measure of attention allocation to the task,[2] although other studies have postulated that cognitive workload better reflects the readiness for resource expenditure.[3] When the cognitive workload required by the task is lower than the available cognitive resources, the task has the potential to be executed successfully. When the cognitive workload imposed by the task exceeds the available resources, task performance is expected to decrease.[3] Older age and age-related neurodegeneration may affect the availability of cognitive resources. With fewer resources available to attend to the task, older adults may show greater workload on a task compared to younger individuals.[3] This increased cognitive workload may reflect inefficient or compensatory use of neural resources to cope with the demand of the task. Some studies have suggested that this increased cognitive workload may serve as a predictor of cognitive decline.[4]

Several techniques have been developed to measure cognitive workload, including questionnaires, performance outcomes, or physiological measures. The National Aeronautics and Space Administration – Task Load Index (NASA-TLX)[5] is one of the most widely used questionnaires of cognitive workload.[6] This questionnaire relies on self-recall of cognitive workload and is typically administered after completion of the task. The NASA-TLX therefore does not provide continuous data but relies on participant’s memory and self-recall of events that have already occurred. Although the psychometric properties of the NASA-TLX have been established in a variety of disciplines such as aviation, military, driving, or skill acquisition, [7 8] the reliability and validity of this instrument have not been tested in older adults with different levels of cognitive functioning.

Performance measures such as accuracy and response times are considered indirect measures of cognitive workload expenditure because they do not directly capture brain activity. A previous study by our group found accuracy and response times on the n-back test to be highly reliable performance measures of working memory in older adults.[9] Unlike performance measures, physiological measures can provide a continuous recording of brain activity in real time. Some studies have suggested that physiological changes may appear before manifestation of symptoms in performance measures, thus providing a more sensitive measure of early cognitive decline.[10-12] In a systematic review, Ranchet et al (2017) scrutinized the physiological changes resulting from increased cognitive workload in older adults with and without cognitive impairment. Increased hemodynamic and electrophysiological activity in the brain, smaller changes in systolic blood pressure, and increased pupillary dilation were observed in healthy older adults compared to younger adults, suggesting additional recruitment of neural resources to cope with task demand. In adults with neurodegenerative conditions, the inability to cope with task demand was even more apparent, resulting in not only an increase in hemodynamic, electrophysiological, and pupillary responses, but also worsening on performance measures.[4] Of those, the pupillary response is particularly interesting since it has been implicated with early tau accumulation in the locus coeruleus (LC) in Alzheimer’s disease (AD).[13] Decreased neuronal density of the LC has been associated with cognitive decline in older adults, mild cognitive impairment, and AD.[14] The LC plays an essential role in the regulation of physiological arousal and cognition.[15] When activated, the LC sends inhibitory projections to the parasympathetic Edinger-Westphal nucleus, which in turn inhibits contraction of the pupillary sphincter muscle.[16] LC activity also triggers the sympathetic nervous system, resulting in activation of the pupillary dilator muscle.[17] A previous study found increased pupillary dilation in participants with single-domain mild cognitive impairment compared to cognitively normal participants, despite performance in the normal ranges.[12] Furthermore, participants with a genetic predisposition for AD were more likely to exhibit larger pupillary size in highly cognitive demanding tasks.[18]

Two methods of pupillary response to cognitive workload have been reported. Task-evoked pupillary response (TEPR) compares the averaged raw pupillary diameter after stimulus onset to the averaged baseline pupillary diameter. Using raw pupillary dilation as a measure of cognitive workload poses some challenges as the light reflex may confound extracting the TEPR, especially in settings where the lighting of the surrounding environment or the luminosity of the screen cannot be entirely controlled.[19] Changes in camera angle and eye movements may also interfere with raw pupillary recording.[17 20] Nonetheless, previous studies found increased TEPR in individuals with elevated risk of AD.[18 21] An alternative to pupillometric baseline-related difference measures is the moment-to-moment pupillary diameter measurement. The Index of Cognitive Activity (ICA) and the Index of Pupillary Activity (IPA) are two moment-to-moment measures that calculate the rate of change of pupillary diameter rather than the difference between averaged pupillary diameter after and before stimulus onset.[22 23] Both ICA and IPA measures are based on the premise that pupils continuously undergo small fluctuations, even in steady illumination conditions.[24] An increase in abrupt discontinuities in the small oscillatory movements of the pupil reflects increased cognitive workload. These two measures of cognitive workload are claimed to successfully separate the pupillary response to cognitive workload from the light reflex. Furthermore, the ICA claims to be unaffected by changes in eye movements and sampling rate.[25] The ICA in particular has been used to investigate changes in cognitive workload in individuals at risk of cognitive impairment, including Parkinson’s disease, multiple sclerosis, and breast cancer.[4 26-30] Overall, the ICA seems to increase with cognitive demand, regardless of disease condition[27]. In addition, some studies report that individuals with increased risk of cognitive impairment show greater ICA compared to controls.[28 30 31] However, the reliability and validity of ICA during working memory tasks in older individuals have not been established.

There is no gold standard for measuring cognitive workload. We selected the P3 (or P300) event-related potential (ERP) as our criterion measure. The P3 is a positive peak at around 300 ms observed in visual or auditory oddball tasks of working memory. This component is considered a sensitive and reliable measure of cognitive workload, including in older adults with cognitive impairment.[4 9 32-34] In addition, the P3 ERP is assumed to share the same neural origins in the LC as the pupillary response to cognitive workload, making this physiological response particularly suitable as a criterion measurement.[35 36]

The aim of this study was to demonstrate the reliability and convergent validity of the NASA-TLX and the ICA in older adults with a wide range of cognitive ability.

## 2 Materials and Methods

### 2.1 Participants

In this test-retest reliability study, 38 right-handed participants were recruited from the University of Kansas Alzheimer’s Disease Center between 05/03/2018 and 03/10/2020. Participants were included in the study if they (1) signed informed consent; (2) were 65 years of age or older; and (3) able to understand the instructions in English. Exclusion criteria were: (1) currently taking steroids, benzodiazepines, or neuroleptics; (2) history of any substance abuse, (3) history of a psychiatric or neurological disorder other than MCI or AD; and (4) vision problems that cannot be resolved by corrective lenses.

Each participant had previously undergone an amyloid PET scan of the brain. Intravenous florbetapir F-18A was administered in a GE Discovery ST-16 PET/CT scanner to assess cerebral amyloid burden. Standard Uptake Value Ratio for six regions of interest was calculated using MIMneuro software (MiM Software Inc, Cleveland, OH) by normalizing the Aβ PET image to the entire cerebellum. Participants were categorized in one of three groups: (1) non-elevated or Aβ-; (2) elevated or Aβ+; or (3) MCI / AD. The recommendations from NIA and the Alzheimer’s Association workgroup were used to categorize participants into Aβ- and Aβ+.[37] The protocol for determination of amyloid elevation is described elsewhere.[38] The average (standard deviation) time between administration of PET scan and pupillometry/EEG assessment was 1090 (479) days. Sixteen were cognitively normal older adults with no elevated amyloid PET scans (Aβ-), 16 were cognitive normal with elevated amyloid PET scans (Aβ+), and six had a clinical diagnosis of MCI or AD. Participants completed their two-week follow-up session 16 ± 7 days after the first session. Each session lasted about 60 minutes including rest breaks.

### 2.2 Procedure

#### 2.2.1 Demographic and Clinical Information

Age, sex, and education were recorded. General cognitive functions were evaluated with the Montreal Cognitive Assessment (MOCA).[39] Scores on the MOCA ranged between 0 and 30.

#### 2.2.2 N-back Test

In this study, the 0-back, 1-back, and 2-back tests were administered. The 0-back test is essentially a memory search task of sustained attention and often used as a control condition.[40 41]. Participants were instructed to press the button as soon as the letter “X” amongst a series of distracter letters appeared on the screen. The 1-back test requires the participant to passively store and update information in working memory. In this test, participants had to press the button if the current letter was the same as the previous letter. The 2-back test requires continuous mental effort to update information of new stimuli and maintain representations of recently presented stimuli in short-term memory.[42] Participants were instructed to press the button when the current letter was the same as the letter presented 2 places before.

Extensive description of the 7-minutes test is provided elsewhere.[9] In short, each n-back test comprised 180 trials, including 60 (33.3%) target trials and 120 (66.7%) nontarget trials. Display time of each letter was 500 ms, followed by a blank interstimulus interval of 1700 ms with a random jitter of 50 ms. Maximum response time was 2150 ms. The participants practiced before the task.

#### 2.2.3 National Aeronautics and Space Administration Task Load Index

The NASA-TLX is one of the most used self-reported questionnaires of cognitive workload. Six items of mental demand, physical demand, temporal demand, effort, performance, and frustration provide a comprehensive measure of cognitive workload.[43] Each item is scored on a visual analogue scale ranging from 0 to 100 in 5-point increments. NASA-TLX was administered immediately after each n-back test. The mean score of the six subscales was computed for each of the conditions and for each subject. In contrast to the original calculation,[5] we did not attribute weights to each of the components since the unweighted average produced better sensitivity and reliability than the weighted average.[44]

#### 2.2.4 Index of Cognitive Activity

While doing the n-back test, participants wore mobile eye tracking glasses (SMI ETG 2, Sensomotoric Instruments, Teltow, Germany). Pupillary size was recorded in real-time at 60 Hz using infrared cameras for both the left and right eye. Pupillary data were analyzed using Eyeworks (Eye Tracking, Inc, Solana Beach, CA, USA). The software analyzed the change in pupil size for each eye throughout each n-back test. Potential artifacts from lighting and eye movements were minimized by using constant room lighting and having the participants focus on the screen. However, even under constant lighting conditions, the pupil continues to oscillate irregularly. Therefore, we transformed raw pupil data to Index of Cognitive Activity (ICA) scores. The ICA discriminates rapid, small bursts in pupillary dilation due to cognitive workload from slower, larger amplitude changes in pupillary size due to the light reflex by decomposing the raw pupillary size to different wavelets of high and low frequency components of the signal. The ICA has a low autocorrelation at a lag of 100 ms, and almost no autocorrelation at a lag of 200 ms. The short latency features and low autocorrelation make the ICA particularly suitable for oddball tasks.[45] The ICA is calculated by dividing the number of rapid small pupillary dilations per second by the number of expected rapid pupillary dilations per second. The values are then transformed using the hyperbolic tangent function. Blinks are factored out by linear interpolation of adjacent time spans to produce continuous values ranging between 0 and 1.

The average percentage of missing data collected from the eye tracker ranged between 0.87% and 2.24%. Three participants had more than 50% of missing ICA values in one or more tests. These values were excluded from the analyses. Mean ICA of the left and right eye were included as outcome measures.

#### 2.2.5 P3 Event-Related Potential

Continuous electro-encephalogram (EEG) was recorded at 1,000 Hz using an Electrical Geodesics high-density system (Magstim EGI, Eugene, OR, USA) with 256 scalp electrodes. The start and end of the task were time-stamped and synchronized with EEG and ICA recordings. EEG recordings were filtered from 0.50 to 30 Hz using EGI software. All other EEG processing was done in EEGLab[46] and in ERPLab.[47] EEG data were online referenced to Cz and offline re-referenced to average of mastoids. Cz was interpolated using the surrounding five channels. Independent component analysis was employed to separate brain activity from ocular, muscular, or cardiovascular artifacts. Signals from bad electrodes were removed and interpolated with the data of surrounding electrodes. Continuous EEG data were segmented into epochs ranging between −100 and 1000 ms of stimulus onset. Each epoch was baseline corrected using the prestimulus interval. Scalp locations and measurement windows for the P3 ERP were based on their spatial extent and latency after inspection of grand average waveform of the task effect. The task effect was calculated by subtracting the average ERP elicited from the targets from the average ERP elicited by non-targets for each participant. The P3 component time window was established between 200 ms and 400 ms for all three tests. Because of the prefrontal cortex involvement in working memory, we identified *a priori* Fz as the main channel, but also report results of the midline electrodes Cz and Pz. No participants were removed from the analyses because of artifacts. P3 peak amplitude of the task effect was considered the main outcome measure to test convergent validity against, but we also calculated P3 peak latency. P3 peak amplitude, and to a lesser extent P3 peak latency, are reliable measures of cognitive workload.[9]

## Data Analysis

Descriptive analysis including mean (standard deviation, SD) and frequency count of participants’ general, performance measures, NASA-TLX, ICA, and ERP data were performed as appropriate. Intra-class correlation coefficients (ICC) were used to calculate test-retest reliability of ICA values and NASA-TLX scores. ICCs were computed as the between subject variance divided by the total (between + within) variance.[48] ICC values less than 0.40 were considered poor; values between 0.40 and 0.59 fair, values between 0.60 and 0.74 good, and values between 0.75 and 1.00 excellent.[49]. Bland-Altman plots were used to visualize the measurement precision of ICA values and NASA-TLX scores across the test moments.[50] Intersubject stability according to subject rankings was calculated using the Pearson r correlation coefficient. Minimal Detectable Change at a 90% confidence interval (MDCR90R) provides an clinically useful indication of absolute reliability and reflects whether an observed change score is above that expected due to measurement error.[51] MDCR90R was calculated as 1.645 x standard error of measurement 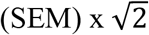 where 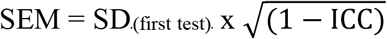. The Kolmogorov–Smirnov test was employed to test the normality of our data distribution in addition to visualization of Q-Q plots. All analyses were done using SAS Enterprise 8.2 and SAS 9.4 software. The threshold of significance was set at α = 0.05.

## 3 Results

### 3.1 Participant Characteristics

Participants (n = 38; 23 (61%) women) were on average 73.81 (5.23) years old and scored 26.97 (2.91) the MOCA scale. MOCA scores of participants ranged between 17 and 30.

### 3.2 Test-Retest Reliability of NASA

Overall, the NASA-TLX scores showed great consistency across the two test moments. ICC scores ranged between 0.71 for 2-back and 0.81 for 0-back, demonstrating good to excellent reliability (Table 1). Pearson r correlations ranged between 0.55 for 2-back and 0.68 for 0-back, indicating strong intersubject stability. MDC of the NASA-TLX ranged from 15.82 points on the 0-back test to 24.33 points on the 2-back.

**Table 1.** Comparison of NASA-TLX and ICA at baseline and two-week follow-up (n = 38).

| Variable | Baseline | Follow-up | Pearson r | ICC, (95% CI) | MDC<sub>90</sub> |
| --- | --- | --- | --- | --- | --- |
| 0-back, NASA-TLX | 19.51 (15.95) | 21.98 (17.89) | 0.68<sup>a</sup> | 0.81 (0.61 – 0.90)<sup>a</sup> | 15.82 |
| 0-back, mean ICA L | 0.33 (0.14) | 0.24 (0.15) | 0.46<sup>b</sup> | 0.63 (0.26 – 0.83)<sup>b</sup> | 0.20 |
| 0-back, mean ICA R | 0.27 (0.17) | 0.28 (0.16) | 0.55<sup>b</sup> | 0.70 (0.40 – 0.85)<sup>a</sup> | 0.22 |
| 1-back, NASA-TLX | 28.24 (17.80) | 27.22 (16.96) | 0.60<sup>a</sup> | 0.78 (0.57 – 0.89)<sup>a</sup> | 19.37 |
| 1-back, mean ICA L | 0.29 (0.17) | 0.27 (0.14) | 0.58<sup>a</sup> | 0.73 (0.47 – 0.86)<sup>a</sup> | 0.20 |
| 1-back, mean ICA R | 0.25 (0.16) | 0.30 (0.15) | 0.39<sup>c</sup> | 0.56 (0.12 – 0.78)<sup>b</sup> | 0.25 |
| 2-back, NASA-TLX | 50.92 (19.41) | 50.61 (19.13) | 0.55<sup>b</sup> | 0.71 (0.42 – 0.85)<sup>a</sup> | 24.33 |
| 2-back, mean ICA L | 0.25 (0.14) | 0.23 (0.16) | 0.50<sup>b</sup> | 0.64 (0.29 – 0.82)<sup>a</sup> | 0.24 |
| 2-back, mean ICA R | 0.24 (0.18) | 0.25 (0.15) | 0.45<sup>b</sup> | 0.62 (0.20 – 0.82)<sup>b</sup> | 0.25 |

Abbreviations: CI, confidence interval; ICA, Index of Cognitive Activity; ICC, Intraclass correlation coefficient; L, left, NASA-TLX, MDC, minimal detectable difference; NASA Task Load Index; R, right.
<sup>a</sup>p<0.0001
<sup>b</sup>p<0.01
<sup>c</sup>p<0.05

Bland-Altman plots showed equal spread of data around the mean (Figure 1). However, the spread of NASA-TLX difference scores (limits of agreement, LOA) was slightly larger in the 2-back test (95% confidence interval (CI), −43.88 – 40.63) compared to 0-back (95% CI, −28.37 – 31.28) and 1-back (95% CI, −31.78 – 30.95) tests.

**Figure 1.**
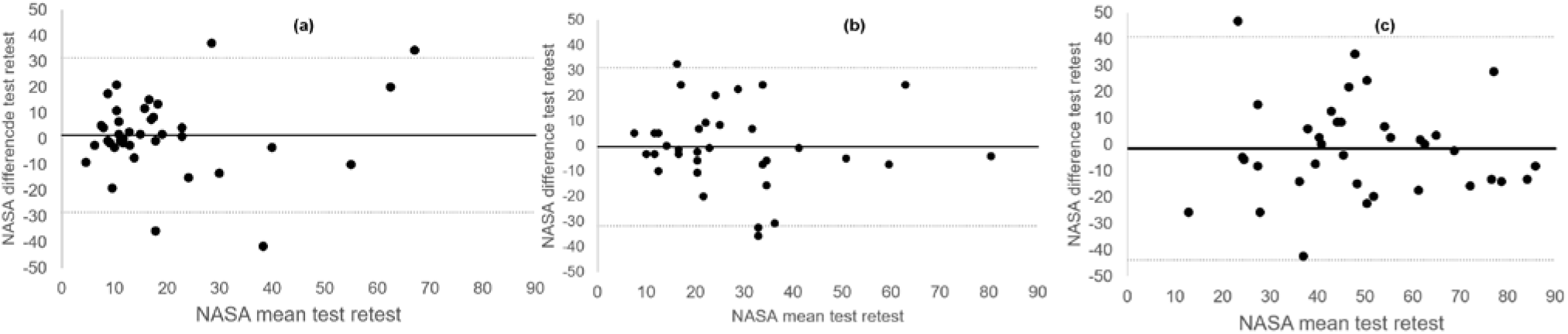
Bland Altman Plots of (a) 0-back NASA-TLX; (b) 1-back NASA-TLX; (c) 2-back NASA-TLX.

### 3.3 Test-Retest Reliability of ICA

All ICC values of the ICA measure were statistically significant (Table 1). ICC values ranged from 0.46 for mean ICA right eye in the 1-back test to 0.73 for mean ICA left eye in the 1-back test. All ICC values produced fair to good reliability. Pearson r correlations ranged from 0.39 (mean ICA right eye in the 1-back test) to 0.58 (mean ICA left eye in 1-back). MDC of ICA values ranged from 0.20 (in 0-back and 1-back) to 0.25 in 2-back for the right eye.

Figure 2 shows the Bland-Altman plots for mean ICA in the left eye for each test. Plot (a) showed a slight tendency towards practice effect in the mean ICA of the left eye during the 0-back test, with decreased ICA at 2-week follow-up compared to baseline assessment. Plots (b) to (f) demonstrated equal distribution of the data around zero, indicating no bias in the results and no heteroscedasticity within the data.

**Figure 2.**
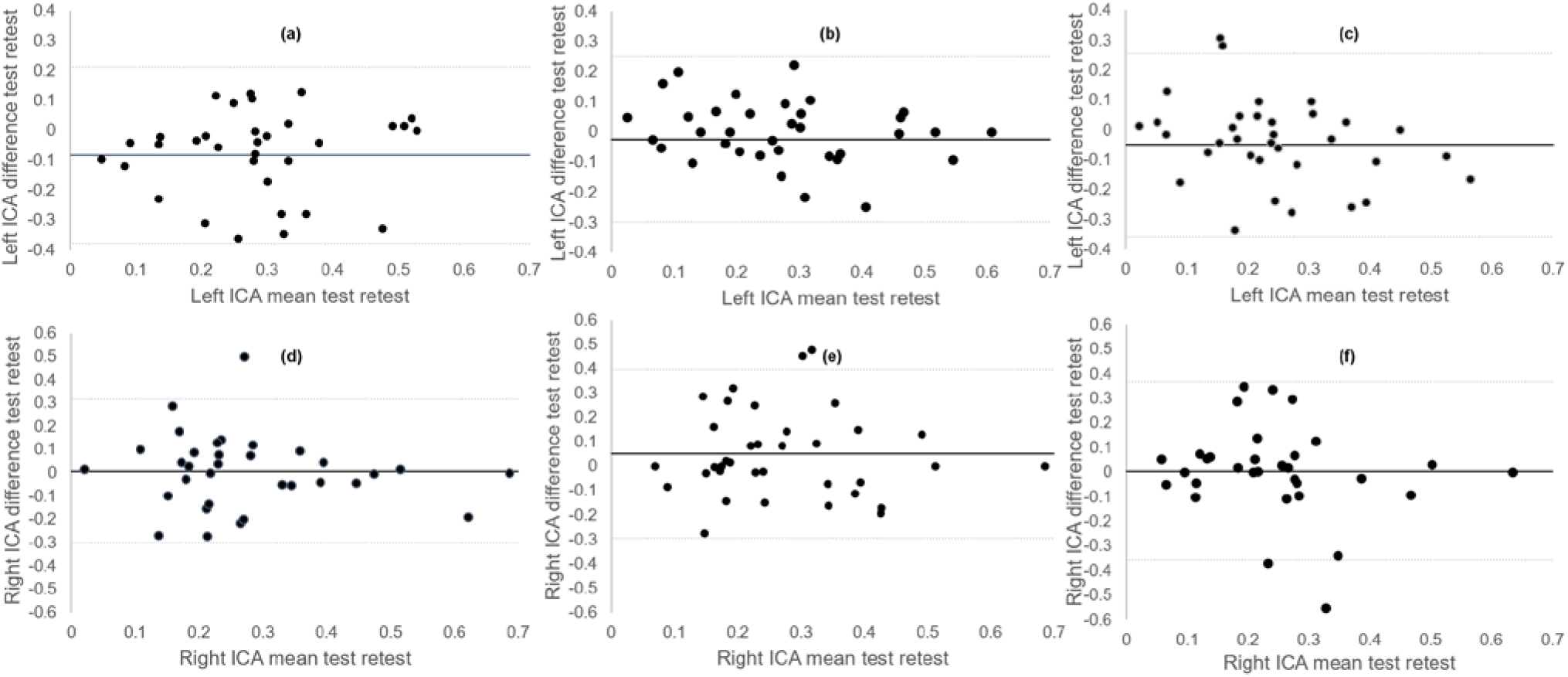
Bland Altman Plots of (a) 0-back ICA mean left eye (b) 1-back ICA mean left eye; (c) 2-back ICA mean left eye; (d) 0-back ICA mean right eye; (e) 1-back ICA mean right eye; (f) 2-back ICA mean right eye. ICA, Index of Cognitive Activity.

### 3.4 Convergent validity of NASA-TLX

There was a trend that higher scores on the NASA-TLX correlated with increased peak P3 latency at channel Fz (r = 0.31; p = 0.06) for the 0-back.

Similar results were found for the 1-back test. Higher NASA-TLX scores correlated with increased peak P3 latency at Pz (r = 0.32; p = 0.05).

No correlations were found between NASA and ERP measures for the 2-back.

### 3.5 Convergent validity of ICA

ICA (mean of left and right eyes) were correlated to ERP (amplitude and latency of Fz, Cz, and Pz). No significant correlations were found between ICA and ERP in 0-back.

Larger mean ICA in the right eye correlated significantly with increased P3 peak latency at Fz (r = 0.32; p = 0.049) and at Cz (r = 0.33; p = 0.048) in the 1-back test. Likewise, larger mean ICA in the left eye correlated with increased P3 peak latency at Pz (r = 0.35; p = 0.03), and with larger P3 peak amplitude at Cz (r = 0.32; p = 0.048).

Larger mean ICA of the left eye correlated with increased P3 peak latency in Pz (r = −0.34; p = 0.04) in the 2-back.

## 4 Discussion

Our results showed that pupillary response, transformed to an Index of Cognitive Activity (ICA), provides fair to good test-retest reliability as a measure of real-time cognitive workload in older adults with and without cognitive impairments. Subjective measures, such as the NASA-TLX, offers even better reliability of cognitive workload in older adults. Moderate correlations were found between these two measures and the P3 ERP.

The ease of use of the NASA-TLX has resulted in applications in diverse fields of aviation, military, human-machine interaction, driving, and medicine.[7] Despite the vast literature, few studies have reported on the test-retest reliability of the NASA-TLX in healthy adults and none in older adults with or without cognitive impairments. Battiste and Bortolussi reported strong test-retest reliability (r = 0.77) of the NASA-TLX in airborne pilots. Hart and Staveland found a correlation of 0.83 in NASA scores administered at baseline and four week follow-up assessment in healthy adults.[5] Xiao reported a test-retest reliability of 0.75 in mental health workers.[52] These correlations coefficients are slightly higher than those found in our study (ranging between 0.55 and 0.68), which may be because our study focused on older adults with a wide range of cognitive ability. Previous studies have shown a potential confounding effect of cognitive impairment on reliability of EEG ERP.[9 53] However, Pearson correlation coefficients tend to overestimate the true test-retest reliability. We extended the correlation analyses with intra-class correlation coefficients (ICC), Bland-Altman plots, and minimal detectable change (MDC) calculations. ICC values provide a single measure of the magnitude of agreement while accounting for the differences in test moments along with the correlation between test moments. The ICC’s showed good to excellent reliability for the NASA-TLX and no signs of systematic bias across the two test moments. The ICC’s were higher and the range scores were smaller (0.71 – 0.81) than those reported in a previous study (range 0.34 – 0.80) that taxed the mental and physical effort of simulated manufacturing tasks in 24 college engineering students.[44]. None of the aforementioned studies provided a graphical representation of the measurement error across the two test moments. The Bland-Altman graphs showed relatively large limits of agreement, with no evidence of test or practice effect in all three tests. MDC calculations showed changes of 15% to 25% of total NASA-TLX scale scores to represent true change beyond measurement error. Taken together, subjective self-recall of cognitive workload is reliable across the spectrum of cognitive aging and has potential to be used as a measure of mental effort expenditure in this population.

Likewise, pupillometry has been used for over five decades as a measure of cognitive workload in the domains of psychophysiology, cognitive neuroscience, and human factors engineering such as aviation or driving. Only recently has pupillometry received attention in the medical field as a potential marker of disease progression in adults with Alzheimer’s disease, Parkinson’s disease, and breast cancer.[12 18 27 30] This rekindled interest in pupillary response to cognitive workload as a marker of cognitive decline warrants an investigation of its psychometric properties. Overall, the ICA produced fair to good reliability scores, ranging between 0.56 and 0.78. These ICC’s are in the same range as the reliability of our convergent measure, the P3 ERP component.[9] Comparison of reliability with other measures of pupillary response is complicated by the type of extraction (TEPR versus ICA), the type of task, and the population of interest. The closest comparison is the study by Kahya et al, that estimated reliability of ICA during postural demanding tasks in Parkinson’s disease.[54] The ICC’s in that study ranged between 0.74 and 0.93, which are higher than those reported in the current study. However, participants in that study completed the retest within hours of the first test, which may have resulted in less day-to-day variability. The Bland-Altman plots revealed no systematic bias of ICA across test moments. MDC values showed a change between 20% and 25% of the total scale score is needed to produce an effect that cannot be attributed to measurement error. These results suggest that ICA provides a stable measure of cognitive workload during cognitive testing in older adults with and without cognitive impairments.

NASA-TLX and ICA correlated only moderately with P3 ERP. A previous study demonstrated a strong correlation (r = −0.70) between peak ICA values and ERP P3 latency in healthy young adults.[55] Comparison of our results with this study is complicated since different ICA metrics (mean versus peak), ERP measures (amplitude versus peak), and population (older versus younger) were used. In addition, this study used a measure of working memory whereas the other study used a cognitive-motor interference balance task. Although n-back is arguably the most ubiquitous working memory test used in ERP studies across the age spectrum,[40] previous studies have shown that the n-back test hosts an array of control processes, including speed of processing, storage, comparison processes, updating, keeping track, task mixing, task shifting, and resistance to interference.[40 41 56] Therefore, the n-back test is a multi-domain cognitive assessment rather than a single-domain test of working memory. These multidomain processes involved with the n-back test may explain the moderate correlations between the ICA and P3 ERP. An alternative explanation is that cognitive workload represents several dimensions of mental, physical, and temporal demand, along with effort, performance, and frustration. It may be that ICA and P3 ERP measure overlapping, yet distinct constructs of cognitive workload. This assumption should be tested in future studies. It also remains unclear why left and right ICA values produced different correlations. While some studies have suggested a lateralization effect of hemispheric function on pupillary response,[57] in this case, the differences are likely due to measurement error.

To our knowledge, this is the first study evaluating the psychometric properties of a subjective and objective measure of cognitive workload in group of older adults with a heterogeneous profile of cognitive ability. We confirmed the cognitive status of each participant using the MOCA. However, we may have missed participants’ true cognitive status because of the lack of detailed cognitive testing and the large time interval since their PET scan. For example, some participants with preclinical AD may have developed cognitive symptoms since their last PET scan, and some participants with the clinical label of MCI may have converted to AD. In addition, we did not establish reliability of ERP in other cognitive domains known to deteriorate in older age, such as memory and language, and this remains an opportunity for further investigation. Future research should investigate the added value of cognitive workload measures in the diagnosis, monitoring, and treatment of individuals at risk of dementia.

## 5 Conclusion

Our current results show that NASA-TLX and ICA are reliable in older adults with and without cognitive impairment. The lack of strong correlation with P3 ERP measure of cognitive workload may be due to the multidimensionality of the construct. Further research is needed to understand the physiological underpinnings of cognitive workload in older adults before these measures can be considered biomarkers of cognitive decline.

## Data Availability

The raw data supporting the conclusions of this article will be made available by the authors, without undue reservation.

## 7 Tables

## 9 Ethics Statement

The studies involving human participants were reviewed and approved by University of Kansas Medical Center Internal Review Board. The patients/participants provided their written informed consent to participate Each participant received $100 for participating in this study.

## 11 Author Contributions

HD, JB, JM, WMB, and KG conceptualized the study. HD, KL, and KG worked out the EEG data processing steps. HD, PA, and KL administered the tests. HD and JM analysed the data. HD wrote the initial manuscript. JB, KL, PA, JM, WMB, and KG reviewed the manuscript and provided valuable comments.

## 13 Acknowledgments

The authors thank the volunteers for their time and willingness to participate in this research. We are also grateful for the staff at the KU Alzheimer Disease Center.

## References

1. Cowan N. The Magical Mystery Four: How is Working Memory Capacity Limited, and Why? Curr Dir Psychol Sci 2010;19(1):51–57 doi: 10.1177/0963721409359277[published Online First: Epub Date]|.

2. Kahneman D. Attention and effort: Citeseer, 1973.

3. Bruya B, Tang Y-Y. Is Attention Really Effort? Revisiting Daniel Kahneman’s Influential 1973 Book Attention and Effort. Frontiers in Psychology 2018;9 doi: 10.3389/fpsyg.2018.01133[published Online First: Epub Date]|.

4. Ranchet M, Morgan JC, Akinwuntan AE, Devos H. Cognitive workload across the spectrum of cognitive impairments: A systematic review of physiological measures. Neuroscience & Biobehavioral Reviews 2017;80:516–37 doi: 10.1016/j.neubiorev.2017.07.001[published Online First: Epub Date]|.

5. Hart SG, Staveland LE. Development of NASA-TLX (Task Load Index): Results of Empirical and Theoretical Research. In: Hancock PA, Meshkati N, eds. Advances in Psychology: North-Holland, 1988:139–83.

6. Dias RD, Ngo-Howard MC, Boskovski MT, Zenati MA, Yule SJ. Systematic review of measurement tools to assess surgeons’ intraoperative cognitive workload. British Journal of Surgery 2018;105(5):491–501 doi: 10.1002/bjs.10795[published Online First: Epub Date]|.

7. Hart SG. Nasa-Task Load Index (NASA-TLX); 20 Years Later. Proceedings of the Human Factors and Ergonomics Society Annual Meeting 2006;50(9):904–08 doi: 10.1177/154193120605000909[published Online First: Epub Date]|.

8. Tubbs-Cooley HL, Mara CA, Carle AC, Gurses AP. The NASA Task Load Index as a measure of overall workload among neonatal, paediatric and adult intensive care nurses. Intensive and Critical Care Nursing 2018;46:64–69 doi: 38TUhttps://doi.org/10.1016/j.iccn.2018.01.004 [publishedU38T Online First: Epub Date]|.

9. Devos H, Burns J, Ahmadnezhad P, et al. Reliability of P3 Event-Related Potential during Working Memory across the Spectrum of Cognitive Aging. medRxiv 2020:2020.05.27.20109157 doi: 10.1101/2020.05.27.20109157[published Online First: Epub Date]|.

10. Ahmadlou M, Adeli A, Bajo R, Adeli H. Complexity of functional connectivity networks in mild cognitive impairment subjects during a working memory task. Clinical Neurophysiology 2014;125(4):694–702 doi: https://doi.org/10.1016/j.clinph.2013.08.033 [published38T Online First: Epub Date]|.

11. Galluzzi S, Nicosia F, Geroldi C, et al. Cardiac autonomic dysfunction is associated with white matter lesions in patients with mild cognitive impairment. J Gerontol A Biol Sci Med Sci 2009;64(12):1312–5 doi: 10.1093/gerona/glp105[published Online First: Epub Date]|.

12. Granholm EL, Panizzon MS, Elman JA, et al. Pupillary Responses as a Biomarker of Early Risk for Alzheimer’s Disease. Journal of Alzheimer’s Disease 2017;56(4):1419–28 doi: 10.3233/jad-161078[published Online First: Epub Date]|.

13. Braak H, Thal DR, Ghebremedhin E, Del Tredici K. Stages of the Pathologic Process in Alzheimer Disease: Age Categories From 1 to 100 Years. Journal of Neuropathology & Experimental Neurology 2011;70(11):960–69 doi: 10.1097/nen.0b013e318232a379[published Online First: Epub Date]|.

14. Wilson RS, Nag S, Boyle PA, et al. Neural reserve, neuronal density in the locus ceruleus, and cognitive decline. Neurology 2013;80(13):1202–08 doi: 10.1212/wnl.0b013e3182897103[published Online First: Epub Date]|.

15. Chandler DJ, Jensen P, McCall JG, Pickering AE, Schwarz LA, Totah NK. Redefining Noradrenergic Neuromodulation of Behavior: Impacts of a Modular Locus Coeruleus Architecture. The Journal of Neuroscience 2019;39(42):8239–49 doi: 10.1523/jneurosci.1164-19.2019[published Online First: Epub Date]|.

16. Samuels E, Szabadi E. Functional Neuroanatomy of the Noradrenergic Locus Coeruleus: Its Roles in the Regulation of Arousal and Autonomic Function Part I: Principles of Functional Organisation. Current Neuropharmacology 2008;6(3):235–53 doi: 10.2174/157015908785777229[published Online First: Epub Date]|.

17. Beatty J, Lucero-Wagoner B. The pupillary system. Handbook of psychophysiology, 2nd ed. New York, NY, US: Cambridge University Press, 000:142–62.

18. Kremen WS, Panizzon MS, Elman JA, et al. Pupillary dilation responses as a midlife indicator of risk for Alzheimer’s disease: association with Alzheimer’s disease polygenic risk. Neurobiology of Aging 2019;83:114–21 doi: 10.1016/j.neurobiolaging.2019.09.001[published Online First: Epub Date]|.

19. Alnæs D, Sneve MH, Espeseth T, Endestad T, van de Pavert SH, Laeng B. Pupil size signals mental effort deployed during multiple object tracking and predicts brain activity in the dorsal attention network and the locus coeruleus. J Vis 2014;14(4) doi: 10.1167/14.4.1[published Online First: Epub Date]|.

20. Mathur A, Gehrmann J, Atchison DA. Pupil shape as viewed along the horizontal visual field. Journal of Vision 2013;13(6):3–3 doi: 10.1167/13.6.3[published Online First: Epub Date]|.

21. Granholm EL, Panizzon MS, Elman JA, et al. Pupillary Responses as a Biomarker of Early Risk for Alzheimer’s Disease. J Alzheimers Dis 2017;56(4):1419–28 doi: 10.3233/jad-161078[published Online First: Epub Date]|.

22. Marshall SP. The Index of Cognitive Activity: measuring cognitive workload. Proceedings of the IEEE 7th Conference on Human Factors and Power Plants 2002:7–7

23. The Index of Pupillary Activity 2018. ACM Press.

24. Stark L, Campbell FW, Atwood J. Pupil Unrest: An Example of Noise in a Biological Servomechanism. Nature 1958;182(4639):857–58 doi: 10.1038/182857a0[published Online First: Epub Date]|.

25. Demberg V, Sayeed A. The Frequency of Rapid Pupil Dilations as a Measure of Linguistic Processing Difficulty. PLOS ONE 2016;11(1):e0146194 doi: 10.1371/journal.pone.0146194[published Online First: Epub Date]|.

26. Devos H, Akinwuntan AE, Alissa N, Morohunfola B, Lynch S. Cognitive performance and cognitive workload in multiple sclerosis: Two different constructs of cognitive functioning? Mult Scler Relat Disord 2020;38:101505 doi: 10.1016/j.msard.2019.101505[published Online First: Epub Date]|.

27. Kahya M, Moon S, Lyons KE, Pahwa R, Akinwuntan AE, Devos H. Pupillary Response to Cognitive Demand in Parkinson’s Disease: A Pilot Study. Front Aging Neurosci 2018;10:90 doi: 10.3389/fnagi.2018.00090[published Online First: Epub Date]|.

28. Moon S, Kahya M, Lyons KE, Pahwa R, Akinwuntan AE, Devos H. Cognitive workload during verbal abstract reasoning in Parkinson’s disease: a pilot study. Int J Neurosci 2020:1–7 doi: 10.1080/00207454.2020.1746309[published Online First: Epub Date]|.

29. Myers JS, Alissa N, Mitchell M, et al. Pilot Feasibility Study Examining Pupillary Response During Driving Simulation as a Measure of Cognitive Load in Breast Cancer Survivors. Oncol Nurs Forum 2020;47(2):203–12 doi: 10.1188/20.Onf.203-212[published Online First: Epub Date]|.

30. Myers JS, Kahya M, Mitchell M, et al. Pupillary response: cognitive effort for breast cancer survivors. Support Care Cancer 2019;27(3):1121–28 doi: 10.1007/s00520-018-4401-0[published Online First: Epub Date]|.

31. Ranchet M, Orlosky J, Morgan J, Qadir S, Akinwuntan AE, Devos H. Pupillary response to cognitive workload during saccadic tasks in Parkinson’s disease. Behav Brain Res 2017;327:162–66 doi: 10.1016/j.bbr.2017.03.043[published Online First: Epub Date]|.

32. Wang C, Gao J, Li M, et al. Association of cognitive impairment and mood disorder with event-related potential P300 in patients with cerebral small vessel diseases. Neuro Endocrinol Lett 2019;40(7-8):333–41

33. Jervis BW, Bigan C, Jervis MW, Besleaga M. New-Onset Alzheimer’s Disease and Normal Subjects 100% Differentiated by P300. American Journal of Alzheimer’s Disease & Other Dementias® 2019;34(5):308–13 doi: 10.1177/1533317519828101[published Online First: Epub Date]|.

34. Ghani U, Signal N, Niazi IK, Taylor D. ERP based measures of cognitive workload: A review. Neuroscience & Biobehavioral Reviews 2020;118:18–26 doi: 10.1016/j.neubiorev.2020.07.020[published Online First: Epub Date]|.

35. Nieuwenhuis S, De Geus EJ, Aston-Jones G. The anatomical and functional relationship between the P3 and autonomic components of the orienting response. Psychophysiology 2011;48(2):162–75 doi: 10.1111/j.1469-8986.2010.01057.x[published Online First: Epub Date]|.

36. Murphy PR, Robertson IH, Balsters JH, O’Connell RG. Pupillometry and P3 index the locus coeruleus-noradrenergic arousal function in humans. Psychophysiology 2011;48(11):1532–43 doi: 10.1111/j.1469-8986.2011.01226.x[published Online First: Epub Date]|.

37. Sperling RA, Aisen PS, Beckett LA, et al. Toward defining the preclinical stages of Alzheimer’s disease: recommendations from the National Institute on Aging-Alzheimer’s Association workgroups on diagnostic guidelines for Alzheimer’s disease. Alzheimers Dement 2011;7(3):280–92 doi: 10.1016/j.jalz.2011.03.003[published Online First: Epub Date]|.

38. Vidoni ED, Yeh H-W, Morris JK, et al. Cerebral β-Amyloid Angiopathy Is Associated with Earlier Dementia Onset in Alzheimer’s Disease. Neurodegenerative Diseases 2016;16(3-4):218–24 doi: 10.1159/000441919[published Online First: Epub Date]|.

39. Nasreddine ZS, Phillips NA, Bedirian V, et al. The Montreal Cognitive Assessment, MoCA: A brief screening tool for mild cognitive impairment. J Am Geriatr Soc 2005;53(4):695–99 doi: 10.1111/j.1532-5415.2005.53221.x[published Online First: Epub Date]|.

40. Bopp KL, Verhaeghen P. Aging and n-Back Performance: A Meta-Analysis. The Journals of Gerontology: Series B 2018 doi: 10.1093/geronb/gby024[published Online First: Epub Date]|.

41. Miller KM, Price CC, Okun MS, Montijo H, Bowers D. Is the N-Back Task a Valid Neuropsychological Measure for Assessing Working Memory? Archives of Clinical Neuropsychology 2009;24(7):711–17 doi: 10.1093/arclin/acp063[published Online First: Epub Date]|.

42. Gevins A, Smith ME, McEvoy LK, et al. A cognitive and neurophysiological test of change from an individual’s baseline. Clinical Neurophysiology 2011;122(1):114–20 doi: 10.1016/j.clinph.2010.06.010[published Online First: Epub Date]|.

43. Hart SG, Staveland LE. Development of NASA-TLX (Task Load Index): Results of empirical and theoretical research. Advances in psychology 1988;52:139–83

44. Ikuma LH, Nussbaum MA, Babski-Reeves KL. Reliability of physiological and subjective responses to physical and psychosocial exposures during a simulated manufacturing task. International Journal of Industrial Ergonomics 2009;39(5):813–20 doi: 38Thttps://doi.org/10.1016/j.ergon.2009.02.005 [published38T Online First: Epub Date]|.

45. Vogels J, Demberg V, Kray J. The Index of Cognitive Activity as a Measure of Cognitive Processing Load in Dual Task Settings. Frontiers in Psychology 2018;9(2276) doi: 10.3389/fpsyg.2018.02276[published Online First: Epub Date]|.

46. Delorme A, Makeig S. EEGLAB: an open source toolbox for analysis of single-trial EEG dynamics including independent component analysis. J Neurosci Methods 2004;134(1):9–21 doi: 10.1016/j.jneumeth.2003.10.009[published Online First: Epub Date]|.

47. Lopez-Calderon J, Luck SJ. ERPLAB: an open-source toolbox for the analysis of event-related potentials. Frontiers in Human Neuroscience 2014;8(213) doi: 10.3389/fnhum.2014.00213[published Online First: Epub Date]|.

48. Shrout PE, Fleiss JL. Intraclass correlations: uses in assessing rater reliability. Psychol Bull 1979;86(2):420–8 doi: 10.1037//0033-2909.86.2.420[published Online First: Epub Date]|.

49. Cicchetti DV. Guidelines, criteria, and rules of thumb for evaluating normed and standardized assessment instruments in psychology. Psychological assessment 1994;6(4):284

50. Bland JM, Altman DG. Statistical methods for assessing agreement between two methods of clinical measurement. Lancet (London, England) 1986;1(8476):307–10

51. Donoghue D, Stokes E. How much change is true change? The minimum detectable change of the Berg Balance Scale in elderly people. Journal of Rehabilitation Medicine 2009;41(5):343–46 doi: 10.2340/16501977-0337[published Online First: Epub Date]|.

52. Xiao YM, Wang ZM, Wang MZ, Lan YJ. [The appraisal of reliability and validity of subjective workload assessment technique and NASA-task load index]. Zhonghua Lao Dong Wei Sheng Zhi Ye Bing Za Zhi 2005;23(3):178–81

53. Lew HL, Gray M, Poole JH. Temporal Stability of Auditory Event-Related Potentials in Healthy Individuals and Patients With Traumatic Brain Injury. Journal of Clinical Neurophysiology 2007;24(5):392–97 doi: 10.1097/wnp.0b013e31814a56e3[published Online First: Epub Date]|.

54. Kahya M, Lyons KE, Pahwa R, Akinwuntan AE, He J, Devos H. Reliability and Validity of Pupillary Response during Dual-task Balance in Parkinson’s Disease. Archives of Physical Medicine and Rehabilitation 2020 doi: 10.1016/j.apmr.2020.08.008[published Online First: Epub Date]|.

55. Kahya M, Liao K, Gustafson K, Akinwuntan A, Devos H. Validation of Pupillary Response Against EEG during Dual-Tasking Postural Control. Archives of Physical Medicine and Rehabilitation 2019;100(10):e142 doi: 10.1016/j.apmr.2019.08.434[published Online First: Epub Date]|.

56. Schmiedek F, Li S-C, Lindenberger U. Interference and facilitation in spatial working memory: Age-associated differences in lure effects in the n-back paradigm. Psychology and Aging 2009;24(1):203–10 doi: 10.1037/a0014685[published Online First: Epub Date]|.

57. Kim M, Barrett AM, Heilman KM. Lateral Asymmetries of Pupillary Responses. Cortex 1998;34(5):753–62 doi: 10.1016/s0010-9452(08)70778-0[published Online First: Epub Date]|.

